## Supplementary Table 1 for "Metabolic imaging across scales reveals distinct prostate cancer phenotypes"

**Supplementary Information file including:**

Supplementary Table 1

Extended Data Figures 1-6

| exp_peak | interval_width_da | compound_name |
|---|---|---|
| **87.0088** | 0.00051129959854279 | Pyruvate |
| **89.0246** | 0.00052314529990838 | Lactate |
| **103.0036** | 0.00060529199649295 | Malonate |
| **112.9877** | 0.00066396247220268 | Oxaloacetate |
| **115.0035** | 0.0006758082859335 | Fumarate |
| **117.0194** | 0.00068765446259533 | Succinate |
| **125.0972** | 0.00073512268198073 | Octanoic acid |
| **131.0351** | 0.00077001646214114 | Glutarate |
| **133.0145** | 0.00078164818512505 | Malate |
| **145.0144** | 0.00085216473715377 | alpha-Ketoglutarate |
| **152.9962** | 0.00089906909218485 | Succinate |
| **153.1287** | 0.0008998474800137 | Decanoic acid |
| **166.9755** | 0.00098121705471499 | Phosphoenolpyruvate |
| **173.0093** | 0.0010166739553483 | cis-Aconitate;Citrate |
| **179.0561** | 0.001052207884598 | Glucose |
| **189.004** | 0.0011106654522308 | Oxalosuccinate |
| **191.02** | 0.001122512497858 | Citrate |
| **208.9846** | 0.0025771426214618 | cis-Aconitate |
| **215.0325** | 0.0012636201084888 | Glucose |
| **224.9801** | 0.001322076059239 | Oxalosuccinate |
| **227.2014** | 0.0013351295196173 | Tetradecanoic acid |
| **241.012** | 0.0014162866034297 | Glucose 1-phosphate |
| **246.9398** | 0.0014511207514829 | Glycerate 1,3-biphosphate |
| **253.2173** | 0.0014880098153043 | Palmitoleic acid |
| **281.2482** | 0.0016527310219772 | Oleic acid |
| **283.2654** | 0.0016645846906727 | Octadecanoic acid |
| **284.056** | 0.0016692306908794 | S-Acetyldihydrolipoamide-E |
| **291.2099** | 0.0017112699781592 | Hexadecanoic acid |
| **317.2244** | 0.0018641420160179 | Oleic acid |
| **319.2406** | 0.0018759897287168 | Octadecanoic acid |
| **398.3282** | 0.0023407415183669 | L-Palmitoylcarnitine |

**Supplementary Table 1.** The list of KEGG glycolysis, TCA cycle, and fatty acid biosynthesis metabolites used for DESI-MSI-derived metabolic pathway enrichment analysis in the secondary cohort.
